# Accident and emergency (AE) attendance in England following infection with SARS-CoV-2 Omicron or Delta

**DOI:** 10.1101/2022.05.03.22274602

**Authors:** Daniel J. Grint, Kevin Wing, Hamish P. Gibbs, Stephen JW Evans, Elizabeth Williamson, Krishnan Bhaskaran, Helen I McDonald, Alex J. Walker, David Evans, George Hickman, Rohini Mathur, Anna Schultze, Christopher T Rentsch, John Tazare, Ian J Douglas, Helen J. Curtis, Caroline E Morton, Sebastian Bacon, Simon Davy, Brian MacKenna, Peter Inglesby, Richard Croker, John Parry, Frank Hester, Sam Harper, Nicholas J DeVito, Will Hulme, Chris Bates, Jonathon Cockburn, Amir Mehrkar, Ben Goldacre, Rosalind M. Eggo, Laurie Tomlinson

## Abstract

The SARS-CoV-2 Omicron variant is increasing in prevalence around the world. Accurate estimation of disease severity associated with Omicron is critical for pandemic planning. We found lower risk of accident and emergency (AE) attendance following SARS-CoV-2 infection with Omicron compared to Delta (HR: 0.39 (95% CI: 0.30 – 0.51; P<.0001). For AE attendances that lead to hospital admission, Omicron was associated with an 85% lower hazard compared with Delta (HR: 0.14 (95% CI: 0.09 – 0.24; P<.0001)).

**Conflicts of Interests:** Nothing to declare.

**Funding statement:** This work was supported by the Medical Research Council MR/V015737/1. TPP provided technical expertise and infrastructure within their data centre *pro bono* in the context of a national emergency. Rosalind Eggo is funded by HDR UK (grant: MR/S003975/1), MRC (grant: MC_PC 19065), NIHR (grant: NIHR200908).

## Introduction

The SARS-CoV-2 variant B.1.1.529 (Omicron) was first identified in South Africa in late 2021. Analysis has found that Omicron is more transmissible than the predominant B.1.617.2 variant (Delta) and it has since become the dominant strain throughout the UK.(1) Only a small proportion of Omicron cases are identified by whole-genome sequencing. In PCR assays for SARS-CoV-2 processed by TaqPath lighthouse laboratories, missingness in one spike protein gene target occurs with the Omicron variant, but not the Delta variant. Spike gene target failure (SGTF) is therefore a proxy for Omicron identification, and has been shown to have excellent sensitivity in England over the study period.(1)

Working on behalf of NHS England, we estimate the risk of accident and emergency (AE) attendance following confirmation of SARS-CoV-2 infection in England, comparing infection with Omicron to Delta, after accounting for demographic factors and comorbidities (Supplement 3).

### Study platform and population

All data were linked, stored and analysed securely within the OpenSAFELY platform https://opensafely.org/ (Supplement 1). The OpenSAFELY dataset is based on 24 million people currently registered with GP surgeries using TPP SystmOne software, covering 40% of England’s population. Pseudonymized data include coded diagnoses, medications and physiological parameters. All code is shared openly for review and re-use under MIT open license (https://github.com/opensafely/SGTF-Omi).

We used linked GP, SARS-CoV-2 testing, vaccination and emergency care data (Supplement 2) to define the study cohort of people first testing positive for SARS-CoV-2 between 5^th^ December 2021 and 1^st^ January 2022. The study was analysed according to the pre-define study protocol (https://github.com/opensafely/SGTF-Omi-research/tree/main/docs), in line with previous work.(2, 3)

SGTF status was known for 330,380/755,432 (44%) people with a first confirmed SARS-CoV-2 infection between 5^th^ December 2021 and 1^st^ January 2022 (237,430 Omicron; 92,950 Delta). A total of 660 (341 Omicron; 319 Delta) AE attendances were recorded with SARS-CoV-2 recorded as the patient diagnosis prior to 21^st^ January 2022, when follow-up was administratively censored. The exposure groups were similar in terms of sex, ethnicity, and regional distribution (Table 1, Supplement 4). The median age of the Omicron group was higher (35 years (interquartile range (IQR) 24 – 49)) vs. 32 (11 – 44), with more comorbidities (2+ comorbidities: 2.4% vs. 1.4%). A lower proportion of Omicron cases were unvaccinated (17.1% vs. 43.0%) while a higher proportion had received a booster vaccination (23.2% vs. 5.1%) compared to Delta at the time of diagnosis.

**Table 1.** Summary demographic and clinical characteristics of the study population.

|  | **Total**<br>**N (%)** | **Delta**<br>**N (%)** | **Omicron**<br>**N (%)** |
| --- | --- | --- | --- |
| **Total population** | 330,380 | 92,950 | 237,430 |
| **AE attendances** | 660 (0.2) | 319 (0.3) | 341 (0.1) |
| **Leading to admission** | 190 (0.1) | 125 (0.1) | 65 (0.0) |
| **Time to AE attendance** |  |  |  |
| Days, median (IQR) | 5.0 (2.0-8.0) | 5.0 (3.0-8.0) | 4.0 (2.0-8.0) |
| **Follow-up time** |  |  |  |
| Days, median (IQR) | 29.0 (24.0-36.0) | 39.0 (34.0-43.0) | 26.0 (23.0-31.0) |
| **Epidemiological week of diagnosis** |  |  |  |
| 05Dec-11Dec | 42,691 (12.9) | 39,317 (42.3) | 3,374 (1.4) |
| 12Dec-18Dec | 63,870 (19.3) | 30,881 (33.2) | 32,989 (13.9) |
| 19Dec-25Dec | 96,025 (29.1) | 15,980 (17.2) | 80,045 (33.7) |
| 26Dec-01Jan | 127,794 (38.7) | 6,772 (7.3) | 121,022 (51.0) |
| **Sex** |  |  |  |
| Female | 170,098 (51.5) | 47,799 (51.4) | 122,299 (51.5) |
| **Age group** |  |  |  |
| 0-39 | 199,764 (60.5) | 60,497 (65.1) | 139,267 (58.7) |
| 40-54 | 81,761 (24.7) | 23,262 (25.0) | 58,499 (24.6) |
| 55-64 | 31,750 (9.6) | 6,730 (7.2) | 25,020 (10.5) |
| 65-74 | 11,607 (3.5) | 1,729 (1.9) | 9,878 (4.2) |
| 75-84 | 4,453 (1.3) | 580 (0.6) | 3,873 (1.6) |
| 85+ | 1,045 (0.3) | 152 (0.2) | 893 (0.4) |
| **Categorical number of comorbidities<sup>a</sup>** |  |  |  |
| None | 296,883 (89.9) | 85,970 (92.5) | 210,913 (88.8) |
| One | 26,517 (8.0) | 5,718 (6.2) | 20,799 (8.8) |
| Two or more | 6,980 (2.1) | 1,262 (1.4) | 5,718 (2.4) |
| **SARS-CoV-2 vaccination status at diagnosis** |  |  |  |
| Unvaccinated | 80,499 (24.4) | 40,007 (43.0) | 40,492 (17.1) |
| First dose >14-days prior | 20,486 (6.2) | 6,187 (6.7) | 14,299 (6.0) |
| Second dose >14-days prior | 169,684 (51.4) | 42,027 (45.2) | 127,657 (53.8) |
| Booster | 59,711 (18.1) | 4,729 (5.1) | 54,982 (23.2) |

<sup>a</sup>Comorbidities as defined in Supplement section 3.

Delta diagnoses were more frequent in the first week of the study period, while Omicron diagnoses predominated thereafter. Consequently, median follow-up time was shorter among the Omicron group (26 days (IQR: 23 - 31)) than the Delta group (39 days (34 - 43)) (Figure 1).

**Figure 1.**
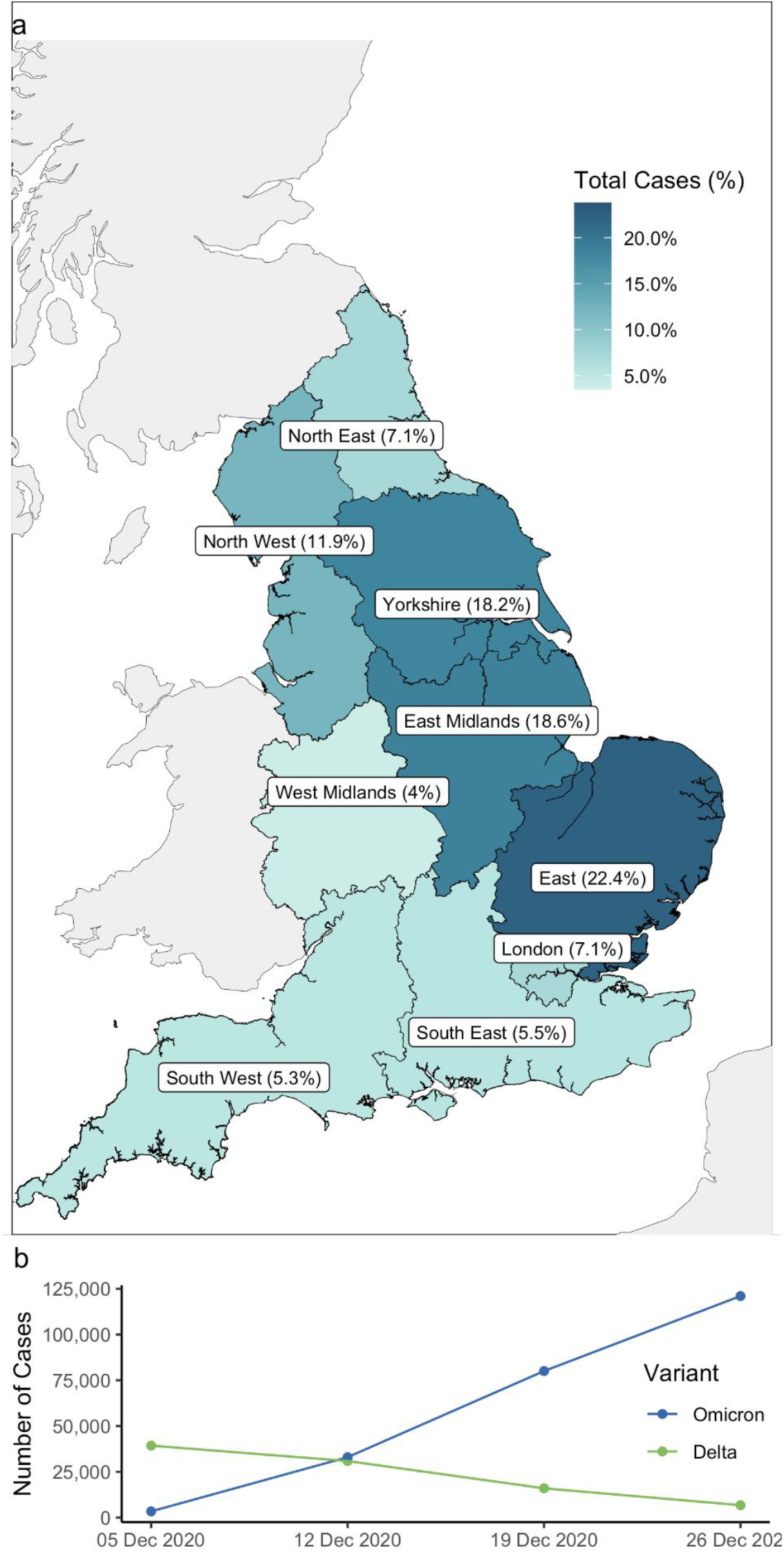
Geographical distribution of study population in England Panel a) Geographical distribution of cohort SARS-CoV-2 diagnoses between 5^th^ December 2021 and 1^st^ January 2022. Panel b) Total number of Omicron and Delta cases by epidemiological week.

### Relative hazard of AE attendance

We estimated the relative hazard of AE attendance with Omicron compared to Delta using Cox proportional hazards regression models stratified by upper tier local authority area (UTLA).(4) Covariate adjustment was informed by a directed acyclic graph (DAG) (Supplement 5). Follow-up began at the date of positive SARS-CoV-2 test and was censored at the earliest of death, AE attendance with diagnosis coded as SARS-CoV-2, or 7-days prior to the emergency care data lock (28^th^ January 2022).

Omicron was consistently associated with lower hazard of AE attendance compared to Delta. In fully-adjusted analysis accounting for demographics, vaccination status, and comorbidities, the hazard of AE attendance was 60% lower for Omicron (hazard ratio (HR): 0.39 (95% confidence interval (CI): 0.30 – 0.51; P <0.0001) compared with Delta.

The hazard of AE attendance was consistently lower for Omicron across all subgroup analyses including epidemiological week, age group, vaccination status, comorbidity status, and ethnicity (Figure 2).

**Figure 2.**
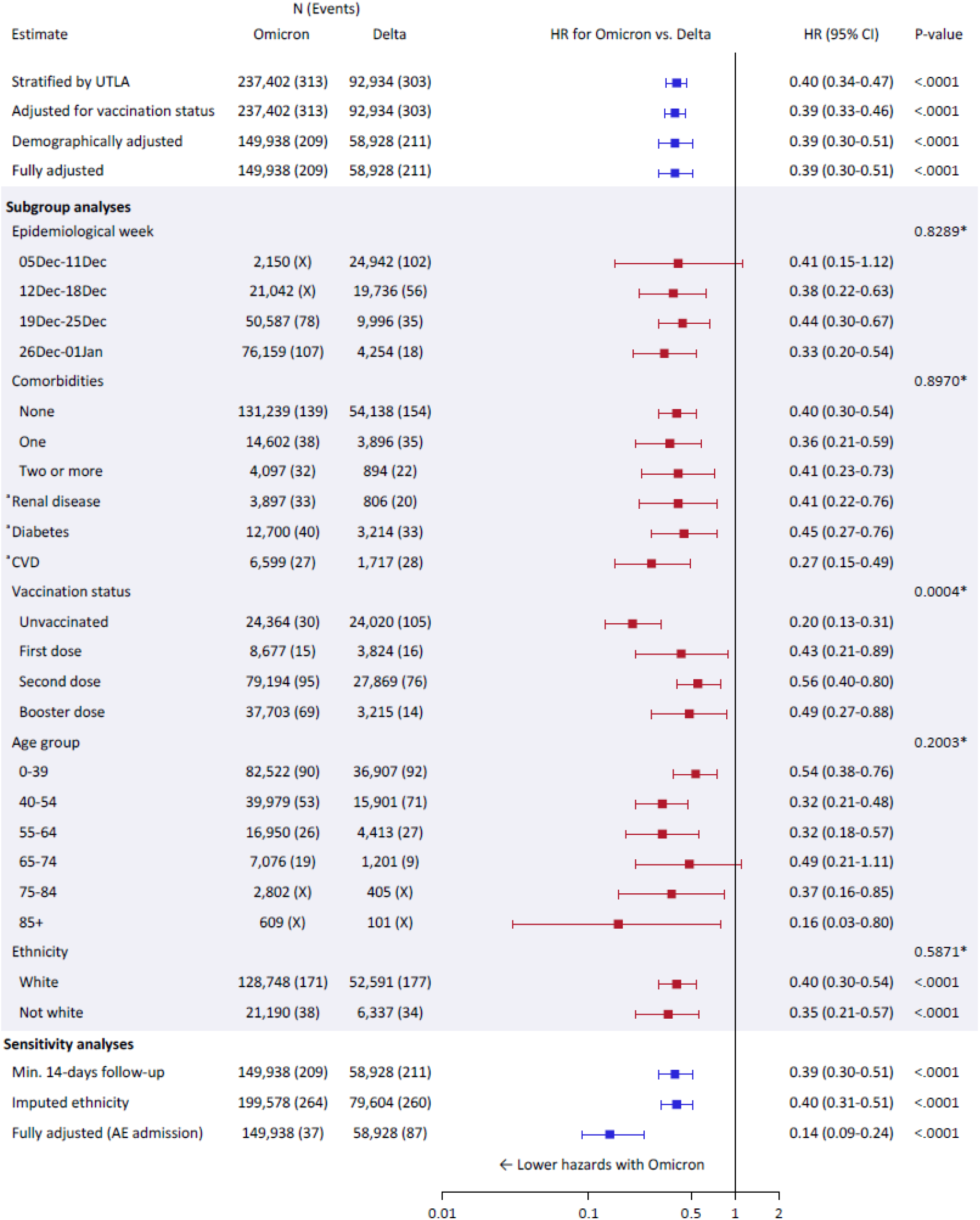
Hazard ratios for AE attendance comparing Omicron vs. Delta from Cox proportional hazards regression stratified by Upper Tier Local Authority (UTLA). All subgroup analyses were performed on the fully-adjusted model.

**Figure 3.**
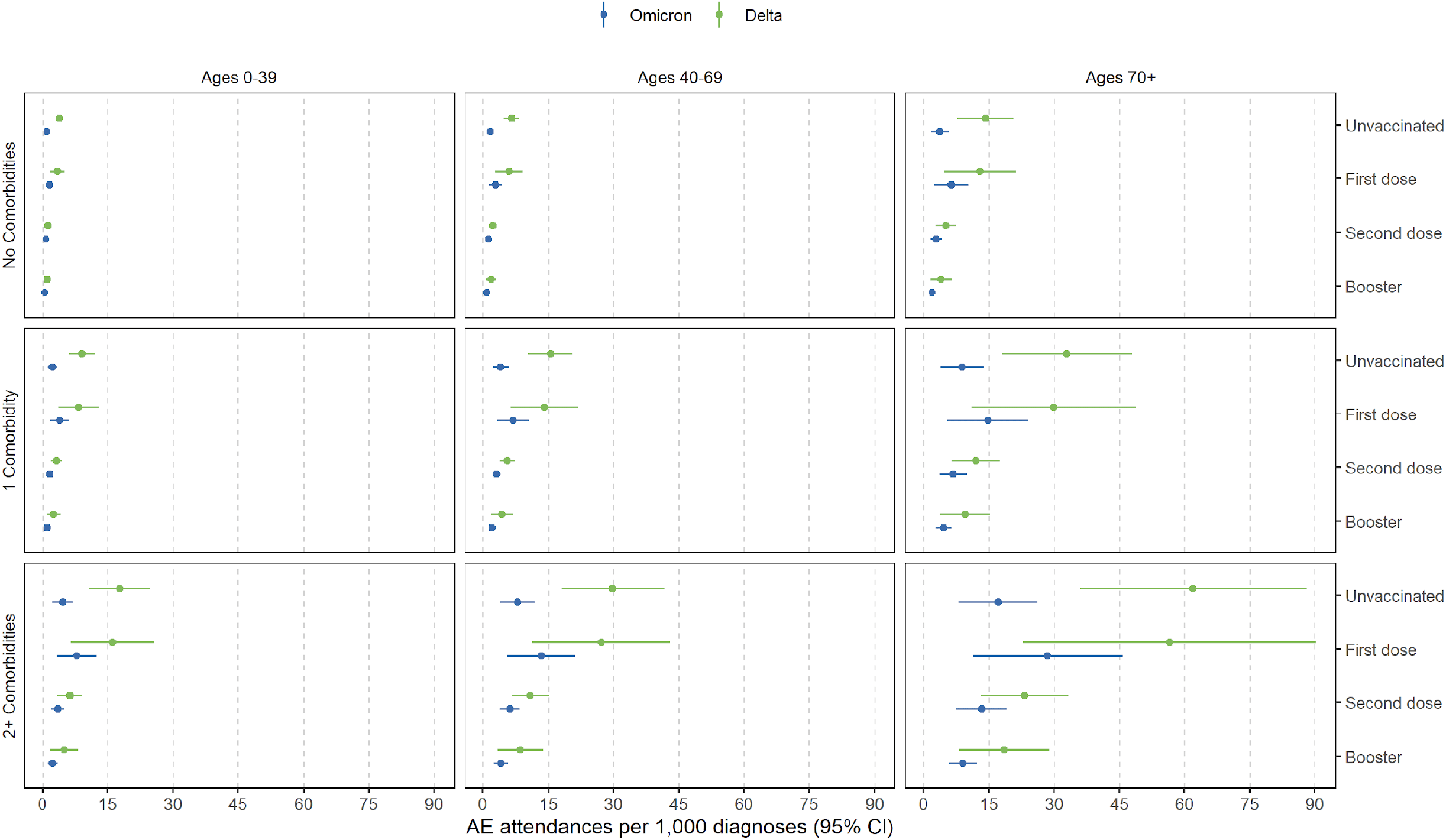
Absolute risk of AE attendance following diagnosis of Omicron or Delta stratified by age, vaccination, and comorbidity status.

There was strong evidence for effect modification by vaccination status (P=.0004). While Omicron was associated with lower hazards of AE attendance regardless of vaccination status, the effect was strongest among the unvaccinated (HR: 0.20 (95% CI: 0.13 – 0.31)) (Figure 2).

44 people who attended AE were excluded from these analyses as they attended AE on the day of testing positive for SARS-CoV-2. In sensitivity analysis, adding one day to all follow-up times to include these outcomes, the relative hazard estimates were unchanged (data not shown). Estimates were also consistent when restricting to people with at least 14-days between testing positive and the data censor, and with multiple imputation for missing ethnicity data (Figure 1).

For AE attendances which resulted in hospital admission, Omicron was associated with an 85% lower hazard compared to Delta (HR: 0.14 (95% CI: 0.09 – 0.24; P<.0001)) (Figure 1).

### Absolute risk of AE attendance

We estimate the absolute risk of AE attendance by 14-days after SARS-CoV-2 positive test by the marginal means from a fully-adjusted logistic regression model, including an interaction term between SARS-CoV-2 variant and vaccination status. This analysis was restricted to positive tests at least 14 days before the censoring date. AE attendances beyond 14 days were censored.

The absolute risk of AE attendance was lower for people double or booster vaccinated for all age and comorbidity subgroups, compared to those unvaccinated. The largest differential in absolute risk of AE attendance between Omicron and Delta was seen for unvaccinated people with two or more comorbidities over the age of 70 (62 AE attendances per 1000 diagnoses (36 - 88) vs. 17 per 1000 (8 - 26)). However, even after booster vaccination, people with two or more comorbidities aged over 70 with Omicron had more than twice the absolute risk of AE attendance compared to unvaccinated people aged over 70 without comorbidities (9 per 1000 (6 - 12) vs. 4 per 1000 (2 - 6)) (Figure 2, Supplement 6).

## Discussion

We show that Omicron is associated with considerably lower risk of AE attendance and in particular admission to hospital following AE attendance than the Delta variant.

The consistency of the effect for all epidemiological weeks shows that the reduced severity with Omicron cannot be explained by other secular changes such as hospitals exceeding capacity or behavioural patterns.

There was strong evidence that the relative reduction in AE attendance for Omicron was largest among the unvaccinated. However, Omicron was consistently associated with a relative reduction in AE attendance regardless of vaccination status and this difference is likely to reflect the greater efficacy of the vaccine against Delta.(5, 6) For the avoidance of doubt, the absolute risk of AE attendance was lower for people double or booster vaccinated for all age and comorbidity subgroups, compared to those unvaccinated.

The relative reduction in AE attendance for Omicron compared with Delta was largest when restricting the outcome to AE attendances which resulted in hospital admission.

Although there was no evidence of differential severity of Omicron compared to Delta by comorbidity status, in the fully-adjusted model those with two or more comorbidities were at greater than 4-fold increased risk of AE attendance compared to those with no reported comorbidities (data not shown). Further, while the absolute risks of AE attendance were consistently lower for Omicron, the risk among people in older age groups living with multiple comorbidities after booster vaccination, remained double that of otherwise healthy unvaccinated older age groups.

This study has several limitations, AE attendance data does not include people who are admitted to hospital without going to AE. Our data include only people first testing positive for SARS-CoV-2. Re- infection with Omicron is common and extra immunity from prior infection may reduce the severity of Omicron further.(7, 8) Our study includes few people in the oldest age groups. Further detailed analysis of which groups remain at greatest risk from Omicron will be essential for health provision planning when societies move toward light touch restrictions in the presence of a high burden of circulating SARS-CoV-2.

## Data Availability

Detailed pseudonymised patient data is potentially re-identifiable and therefore not shared.

## Ethical approval

This study was approved by the Health Research Authority (REC reference 20/LO/0651) and by the LSHTM Ethics Board (reference 21863).

## Acknowledgements

We are grateful for the support received from the TPP Technical Operations team and for generous assistance from the information governance and database teams at NHS England / NHSX.

## Figure 1 legend

^a^Seperate models adjusted for age and vaccination status only. *Likelihood ratio test for interaction between exposure group and subgroup. Number of events marked X are masked to prevent identifiability. All models are stratified on region by UTLA.

Demographically adjusted model includes adjustment for: age, sex, vaccination status, IMD, ethnicity, household size, rural urban classification, epidemiological week, and care home status. The fully-adjusted model includes adjustment for: age, sex, vaccination status, IMD, ethnicity, smoking status, obesity, household size, rural urban classification, comorbidities, epidemiological week, and care home status. There was weak evidence of non-proportional hazards in this model (global test of Schoenfeld residuals, P=0.052). However, this was driven by the cubic spline terms for age, for the primary exposure SGTF there was no evidence of non-proportional hazards (P=0.17).

The first sensitivity analysis is restricted to people with a minimum of 14-days from testing positive for SARS-CoV-2 to the follow-up censor. In the second sensitivity analysis missing data on ethnicity has been imputed using multiple imputation. In the final sensitivity analysis the outcome is defined by admission to hospital following AE attendance.

