## Supplementary material for "Accident and emergency (AE) attendance in England following infection with SARS-CoV-2 Omicron or Delta"

#### Table of Contents

1. Further information on OpenSAFELY 2

2. Details on data sources 3

3. Definition of comorbidities 4

4. Complete demographic and clinical characteristics 6

5. Theoretical causal pathway design, directed acyclic graph (DAG) 8

6. Absolute risk of AE attendance following diagnosis of Omicron and Delta stratified by age, vaccination, and comorbidity status 9

7. References 10

### 1. Further information on OpenSAFELY

All data were linked, stored and analysed securely within the OpenSAFELY platform https://opensafely.org/. The dataset analysed within OpenSAFELY is based on 24 million people currently registered with GP surgeries using TPP SystmOne software. Data include pseudonymized data such as coded diagnoses, medications and physiological parameters. No free text data are included. All code is shared openly for review and re-use under MIT open license (https://github.com/opensafely/SGTF-Omi). Detailed pseudonymised patient data is potentially re-identifiable and therefore not shared. We rapidly delivered the OpenSAFELY data analysis platform without prior funding to deliver timely analyses on urgent research questions in the context of the global Covid-19 health emergency: now that the platform is established we are developing a formal process for external users to request access in collaboration with NHS England; details of this process will be published shortly on OpenSAFELY.org.

#### Information governance and ethics

NHS England is the data controller; TPP is the data processor; and the key researchers on OpenSAFELY are acting on behalf of NHS England. This implementation of OpenSAFELY is hosted within the TPP environment which is accredited to the ISO 27001 information security standard and is NHS IG Toolkit compliant;[1,2] patient data has been pseudonymised for analysis and linkage using industry standard cryptographic hashing techniques; all pseudonymised datasets transmitted for linkage onto OpenSAFELY are encrypted; access to the platform is via a virtual private network (VPN) connection, restricted to a small group of researchers; the researchers hold contracts with NHS England and only access the platform to initiate database queries and statistical models; all database activity is logged; only aggregate statistical outputs leave the platform environment following best practice for anonymisation of results such as statistical disclosure control for low cell counts.[3] The OpenSAFELY research platform adheres to the obligations of the UK General Data Protection Regulation (GDPR) and the Data Protection Act 2018. In March 2020, the Secretary of State for Health and Social Care used powers under the UK Health Service (Control of Patient Information) Regulations 2002 (COPI) to require organisations to process confidential patient information for the purposes of protecting public health, providing healthcare services to the public and monitoring and managing the COVID-19 outbreak and incidents of exposure; this sets aside the requirement for patient consent.[4] Taken together, these provide the legal bases to link patient datasets on the OpenSAFELY platform. GP practices, from which the primary care data are obtained, are required to share relevant health information to support the public health response to the pandemic, and have been informed of the OpenSAFELY analytics platform.

This study was approved by the Health Research Authority (REC reference 20/LO/0651) and by the LSHTM Ethics Board (reference 21863).

#### 2. Details on data sources

We used several linked data sets in this analysis (Table S1).

| Data | Description |
|---|---|
| SARS-CoV-2 tests | Positive and negative tests of people tested under the UK's Pillar 1 and Pillar 2 testing schemes are reported.[5] Pillar 2 generally is community tests and Pillar 1 is tests in hospital (patients and health care workers). Only some of the labs used for testing in England use the 3 channel PCR for which a "failure to detect the Spike-gene target" is indicative of the VOC, therefore not all positive tests have known SGTF status. The data come from Public Health England's (PHE) Second Generation Surveillance System. |
| General Practitioner (GP) data | GP data are drawn from patients who are registered at a practice that runs the TPP SystmOne (https://www.tpp-uk.com/products/systmone). This is approximately 40% of GPs in England. Each patient encounter with a GP is coded using CTV3 codes, which fully aligns with SNOMED-CT which describe the reason for the encounter, and these codes are used to define the health history of each individual.[6] Prescribed medications are also stored in the health record. Demographic data such as age and ethnicity are collected by GPs, as are some behavioural data like whether an individual smokes. |
| Hospital admission | Secondary Uses Statistics (SUS) data used to define hospital admission and ICU admission. Data are collected on hospital admissions with ICD-10 codes for conditions, and procedural codes for treatment. Whether the patient is admitted to intensive care during their hospital admission is also included. (https://digital.nhs.uk/services/secondary-uses-service-sus) |
| Emergency care | The Emergency Care Data Set (ECDS) is the national data set for urgent and emergency care. ECDS is part of Hospital Episode Statistics (HES) and is provided to OpenSAFELY via NHS Digital's Secondary Use Service (SUS). (https://digital.nhs.uk/data-and-information/data-collections-and-data-sets/data-sets/emergency-care-data-set-ecds) |
| Mortality date | The date of death plus codes for the cause of death are from the Office for National Statistics. We only use date of death in this study. |
| Vaccination date | The date, dose number, vaccine manufacturer and batch are entered into their health record. We only use the date of administration of the first dose in this study. |
| Index of multiple deprivation (IMD) | We use the England IMD which is matched to individuals at the postcode level. |
| Urban/Rural classification | We use 5 categories of Urban/Rural classifications which are matched to individuals at the postcode level. |

**Table S1**. Data sources used in this analysis.

### 3. Definition of comorbidities

A patient is identified as having a comorbidity if their health record includes codes indicative of each of the conditions (Table S2).

| **Condition** | **Codelist defining presence of comorbidity** |
| --- | --- |
| Aplastic anaemia | https://codelists.opensafely.org/codelist/opensafely/aplastic-anaemia/ |
| Asplenia | https://codelists.opensafely.org/codelist/opensafely/asplenia/ |
| Asthma | https://codelists.opensafely.org/codelist/opensafely/asthma-diagnosis/<br>https://codelists.opensafely.org/codelist/opensafely/asthma-inhaler-salbutamol-medication/2020-04-15/<br>https://codelists.opensafely.org/codelist/opensafely/asthma-inhaler-steroid-medication/2020-04-15/<br>https://codelists.opensafely.org/codelist/opensafely/asthma-oral-prednisolone-medication/2020-04-27/ |
| Bone marrow transplant | https://codelists.opensafely.org/codelist/opensafely/bone-marrow-transplant/2020-04-15/ |
| Cancer | https://codelists.opensafely.org/codelist/opensafely/cancer-excluding-lung-and-haematological/2020-04-15/<br>https://codelists.opensafely.org/codelist/opensafely/chemotherapy-or-radiotherapy-updated/2020-04-15/<br>https://codelists.opensafely.org/codelist/opensafely/haematological-cancer/2020-04-15/<br>https://codelists.opensafely.org/codelist/opensafely/lung-cancer/2020-04-15/ |
| Chronic cardiac disease | https://codelists.opensafely.org/codelist/opensafely/chronic-cardiac-disease/2020-04-08/ |
| Chronic respiratory disease | https://codelists.opensafely.org/codelist/opensafely/chronic-respiratory-disease/2020-04-10/ |
| Chronic liver disease | https://codelists.opensafely.org/codelist/opensafely/chronic-liver-disease/2020-06-02/ |
| Dementia | https://codelists.opensafely.org/codelist/opensafely/dementia/2020-04-22/ |
| Diabetes | https://codelists.opensafely.org/codelist/opensafely/diabetes/2020-04-15/ |
| Chronic kidney disease | https://codelists.opensafely.org/codelist/opensafely/chronic-kidney-disease/2020-04-14/ |
| GI bleed | https://codelists.opensafely.org/codelist/opensafely/gi-bleed-or-ulcer/2020-04-08/ |
| HIV | https://codelists.opensafely.org/codelist/opensafely/hiv/2020-07-13/ |
| Permanent immunosuppression | https://codelists.opensafely.org/codelist/opensafely/permanent-immunosuppression/2020-06-02/ |
| Temporary immunosuppression | https://codelists.opensafely.org/codelist/opensafely/temporary-immunosuppression/2020-04-24/ |
| Hypertension | https://codelists.opensafely.org/codelist/opensafely/hypertension/2020-04-28/ |
| Stroke | https://codelists.opensafely.org/codelist/opensafely/stroke-updated/2020-06-02/ |
| Inflammatory bowel disease | https://codelists.opensafely.org/codelist/opensafely/inflammatory-bowel-disease/2020-04-07/ |
| Neurological conditions | |

| | |
|---|---|
| Psoriasis<br><br>Sickle cell disease<br><br>Smoking<br><br>Organ transplant | https://codelists.opensafely.org/codelist/opensafely/other-neurological-conditions/2020-06-02/<br>https://codelists.opensafely.org/codelist/opensafely/ra-sle-psoriasis/2020-04-14/<br>https://codelists.opensafely.org/codelist/opensafely/sickle-cell-disease/2020-04-14/<br>https://codelists.opensafely.org/codelist/opensafely/smoking-clear/2020-04-29/<br>https://codelists.opensafely.org/codelist/opensafely/smoking-unclear/2020-04-29/<br>https://codelists.opensafely.org/codelist/opensafely/solid-organ-transplantation/2020-04-10/ |

**Table S2**. Comorbidities and the codelists that determine if a patient is classified as having that comorbidity. All codelists are reviewed by clinicians.

#### 4. Complete demographic and clinical characteristics

| | Total N (%) | Delta N (%) | Omicron N (%) |
|---|---|---|---|
| **Total population** | 330,380 | 92,950 | 237,430 |
| **AE attendances** | 660 (0.2) | 319 (0.3) | 341 (0.1) |
| **Leading to admission** | 190 (0.1) | 125 (0.1) | 65 (0.0) |
| **Time to AE attendance** | | | |
| Days, median (IQR) | 5.0 (2.0-8.0) | 5.0 (3.0-8.0) | 4.0 (2.0-8.0) |
| **Follow-up time** | | | |
| Days, median (IQR) | 29.0 (24.0-36.0) | 39.0 (34.0-43.0) | 26.0 (23.0-31.0) |
| **Epidemiological week of diagnosis** | | | |
| 05Dec-11Dec | 42,691 (12.9) | 39,317 (42.3) | 3,374 (1.4) |
| 12Dec-18Dec | 63,870 (19.3) | 30,881 (33.2) | 32,989 (13.9) |
| 19Dec-25Dec | 96,025 (29.1) | 15,980 (17.2) | 80,045 (33.7) |
| 26Dec-01Jan | 127,794 (38.7) | 6,772 (7.3) | 121,022 (51.0) |
| **Sex** | | | |
| Female | 170,098 (51.5) | 47,799 (51.4) | 122,299 (51.5) |
| **Age** | | | |
| Years, median (IQR) | 34.0 (22.0-48.0) | 32.0 (11.0-44.0) | 35.0 (24.0-49.0) |
| **Age group** | | | |
| 0-39 | 199,764 (60.5) | 60,497 (65.1) | 139,267 (58.7) |
| 40-54 | 81,761 (24.7) | 23,262 (25.0) | 58,499 (24.6) |
| 55-64 | 31,750 (9.6) | 6,730 (7.2) | 25,020 (10.5) |
| 65-74 | 11,607 (3.5) | 1,729 (1.9) | 9,878 (4.2) |
| 75-84 | 4,453 (1.3) | 580 (0.6) | 3,873 (1.6) |
| 85+ | 1,045 (0.3) | 152 (0.2) | 893 (0.4) |
| **SARS-CoV-2 vaccination status at diagnosis** | | | |
| Unvaccinated | 80,499 (24.4) | 40,007 (43.0) | 40,492 (17.1) |
| First dose | 20,486 (6.2) | 6,187 (6.7) | 14,299 (6.0) |
| Second dose | 169,684 (51.4) | 42,027 (45.2) | 127,657 (53.8) |
| Booster | 59,711 (18.1) | 4,729 (5.1) | 54,982 (23.2) |
| **Ethnicity, in 5 categories** | | | |
| White | 213,109 (64.5) | 61,072 (65.7) | 152,037 (64.0) |
| South Asian | 16,112 (4.9) | 3,916 (4.2) | 12,196 (5.1) |
| Black | 7,025 (2.1) | 1,149 (1.2) | 5,876 (2.5) |
| Mixed | 4,551 (1.4) | 1,186 (1.3) | 3,365 (1.4) |
| Other | 4,018 (1.2) | 1,023 (1.1) | 2,995 (1.3) |
| Missing | 85,565 (25.9) | 24,604 (26.5) | 60,961 (25.7) |
| **Evidence of obesity (missing set to none)[a]** | | | |
| No record of obesity | 273,603 (82.8) | 79,587 (85.6) | 194,016 (81.7) |
| Obese I (30-34.9) | 35,543 (10.8) | 8,214 (8.8) | 27,329 (11.5) |
| Obese II (35-39.9) | 14,201 (4.3) | 3,409 (3.7) | 10,792 (4.5) |
| Obese III (40+) | 7,033 (2.1) | 1,740 (1.9) | 5,293 (2.2) |
| **Smoking status (missing set to never)[b]** | | | |
| Never | 206,862 (62.6) | 63,098 (67.9) | 143,764 (60.6) |

| | | | |
|---|---|---|---|
| Former | 83,960 (25.4) | 20,144 (21.7) | 63,816 (26.9) |
| Current | 39,558 (12.0) | 9,708 (10.4) | 29,850 (12.6) |
| **Categorical number of comorbidities** | | | |
| No comorbidity | 296,883 (89.9) | 85,970 (92.5) | 210,913 (88.8) |
| 1 comorbidity | 26,517 (8.0) | 5,718 (6.2) | 20,799 (8.8) |
| 2+ comorbidities | 6,980 (2.1) | 1,262 (1.4) | 5,718 (2.4) |
| **Index of Multiple Deprivation (IMD)** | | | |
| 1 least deprived | 67,072 (20.3) | 19,304 (20.8) | 47,768 (20.1) |
| 2 | 66,028 (20.0) | 18,250 (19.6) | 47,778 (20.1) |
| 3 | 64,845 (19.6) | 17,619 (19.0) | 47,226 (19.9) |
| 4 | 67,320 (20.4) | 18,115 (19.5) | 49,205 (20.7) |
| 5 most deprived | 65,115 (19.7) | 19,662 (21.2) | 45,453 (19.1) |
| **Categorical household size** | | | |
| 1-2 | 86,660 (26.2) | 18,814 (20.2) | 67,846 (28.6) |
| 3-5 | 169,309 (51.2) | 53,271 (57.3) | 116,038 (48.9) |
| 6-10 | 21,739 (6.6) | 7,182 (7.7) | 14,557 (6.1) |
| 11+ | 1,636 (0.5) | 373 (0.4) | 1,263 (0.5) |
| Missing | 51,036 (15.4) | 13,310 (14.3) | 37,726 (15.9) |
| **Binary care home status[c]** | | | |
| Private home | 330,180 (99.9) | 92,914 (100.0) | 237,266 (99.9) |
| Care home | 200 (0.1) | 36 (0.0) | 164 (0.1) |
| **NHS England region** | | | |
| East | 73,990 (22.4) | 22,131 (23.8) | 51,859 (21.8) |
| East Midlands | 61,328 (18.6) | 18,856 (20.3) | 42,472 (17.9) |
| London | 23,374 (7.1) | 3,288 (3.5) | 20,086 (8.5) |
| North East | 23,360 (7.1) | 5,371 (5.8) | 17,989 (7.6) |
| North West | 39,290 (11.9) | 11,581 (12.5) | 27,709 (11.7) |
| South East | 18,062 (5.5) | 4,545 (4.9) | 13,517 (5.7) |
| South West | 17,568 (5.3) | 6,053 (6.5) | 11,515 (4.8) |
| West Midlands | 13,224 (4.0) | 3,853 (4.1) | 9,371 (3.9) |
| Yorkshire and the Humber | 60,034 (18.2) | 17,243 (18.6) | 42,791 (18.0) |
| **Rural Urban in five categories** | | | |
| Urban major conurbation | 76,189 (23.1) | 16,792 (18.1) | 59,397 (25.0) |
| Urban minor conurbation | 28,700 (8.7) | 7,172 (7.7) | 21,528 (9.1) |
| Urban city and town | 164,778 (49.9) | 49,727 (53.5) | 115,051 (48.5) |
| Rural town and fringe | 36,288 (11.0) | 11,325 (12.2) | 24,963 (10.5) |
| Rural village and dispersed | 21,599 (6.5) | 7,105 (7.6) | 14,494 (6.1) |
| Missing | 2,826 (0.9) | 829 (0.9) | 1,997 (0.8) |

[a]120,629 Missing values set to no evidence of obesity; [b]63,542 missing values set to never smoking status; [c]Based on address linkage with CQC data as of 1st Feb 2020: 2,826 missing values set to private home.

#### 5. Theoretical causal pathway design, directed acyclic graph (DAG)

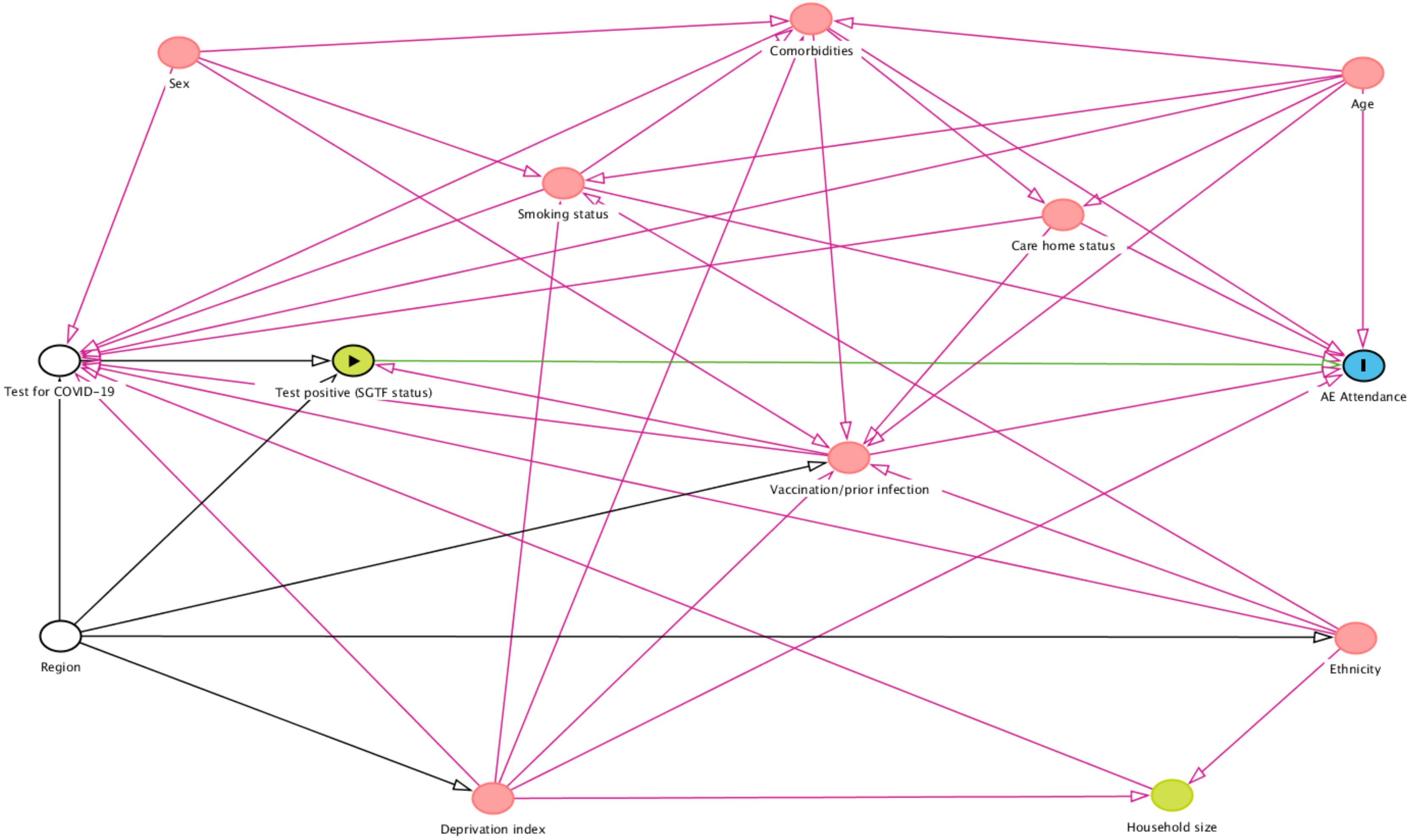

#### 6. Absolute risk of AE attendance following diagnosis of Omicron and Delta stratified by age, vaccination, and comorbidity status

| Comorbidities/Vax/Age group | Delta (95% CI) | Omicron (95% CI) |
|---|---|---|
| **No Comorbidities** | | |
| Unvaccinated: 0-39 | 0.39 (0.31-0.47) | 0.10 (0.06-0.14) |
| 40-69 | 0.66 (0.48-0.84) | 0.17 (0.10-0.24) |
| 70+ | 1.42 (0.78-2.06) | 0.37 (0.17-0.58) |
| First dose: 0-39 | 0.35 (0.18-0.52) | 0.17 (0.09-0.25) |
| 40-69 | 0.60 (0.28-0.91) | 0.29 (0.14-0.44) |
| 70+ | 1.29 (0.47-2.11) | 0.63 (0.23-1.02) |
| Second dose: 0-39 | 0.14 (0.10-0.18) | 0.08 (0.06-0.10) |
| 40-69 | 0.23 (0.17-0.29) | 0.13 (0.10-0.16) |
| 70+ | 0.51 (0.28-0.74) | 0.29 (0.16-0.42) |
| Booster: 0-39 | 0.11 (0.05-0.17) | 0.05 (0.03-0.07) |
| 40-69 | 0.19 (0.08-0.29) | 0.09 (0.06-0.12) |
| 70+ | 0.40 (0.16-0.65) | 0.19 (0.12-0.27) |
| **1 Comorbidity** | | |
| Unvaccinated: 0-39 | 0.92 (0.62-1.21) | 0.24 (0.13-0.34) |
| 40-69 | 1.55 (1.04-2.06) | 0.41 (0.23-0.59) |
| 70+ | 3.29 (1.79-4.79) | 0.88 (0.39-1.37) |
| First dose: 0-39 | 0.83 (0.37-1.29) | 0.40 (0.19-0.62) |
| 40-69 | 1.41 (0.63-2.18) | 0.69 (0.32-1.06) |
| 70+ | 2.99 (1.10-4.88) | 1.48 (0.55-2.41) |
| Second dose: 0-39 | 0.33 (0.20-0.45) | 0.18 (0.12-0.25) |
| 40-69 | 0.56 (0.38-0.73) | 0.31 (0.22-0.41) |
| 70+ | 1.20 (0.64-1.75) | 0.68 (0.37-0.99) |
| Booster: 0-39 | 0.26 (0.10-0.42) | 0.12 (0.08-0.17) |
| 40-69 | 0.44 (0.19-0.69) | 0.21 (0.14-0.28) |
| 70+ | 0.95 (0.38-1.52) | 0.46 (0.28-0.64) |
| **2+ Comorbidities** | | |
| Unvaccinated: 0-39 | 1.78 (1.07-2.48) | 0.47 (0.23-0.70) |
| 40-69 | 2.98 (1.80-4.17) | 0.80 (0.40-1.19) |
| 70+ | 6.20 (3.59-8.82) | 1.71 (0.80-2.62) |
| First dose: 0-39 | 1.61 (0.66-2.57) | 0.79 (0.33-1.25) |
| 40-69 | 2.71 (1.12-4.30) | 1.34 (0.56-2.12) |
| 70+ | 5.66 (2.28-9.03) | 2.85 (1.13-4.57) |
| Second dose: 0-39 | 0.64 (0.35-0.92) | 0.36 (0.21-0.51) |
| 40-69 | 1.08 (0.66-1.51) | 0.62 (0.38-0.85) |
| 70+ | 2.32 (1.31-3.33) | 1.33 (0.75-1.90) |
| Booster: 0-39 | 0.51 (0.18-0.83) | 0.24 (0.13-0.36) |
| 40-69 | 0.86 (0.34-1.38) | 0.42 (0.25-0.58) |
| 70+ | 1.85 (0.81-2.89) | 0.90 (0.58-1.22) |
